# Development and Validation of Novel Cell-free Direct Neutralization Assay for SARS-CoV-2

**DOI:** 10.1101/2024.07.24.24310905

**Authors:** Ji Youn Lim, Alyssa Fiore, Bruce Le, Corinne Minzer, Halle White, Krystle Burinski, Humaira Janwari, David Wright, Sasha Perebikovsky, Ralph Davis, David Okrongly, Aravind Srinivasan

## Abstract

Neutralizing antibody titer elicited through infection or vaccination is accepted as a reliable surrogate for protection from SARS-CoV-2 infection, hospitalization, and mortality. The gold standard for measuring neutralizing antibody levels relies on culturing live virus in the presence of a target cell and quantitating the level where 50% of the target cells are infected. These assays have numerous technical challenges, not the least is the requirement for a BSL-3 laboratory to perform the live virus testing. We developed the Q-NAb IgG Test for the quantitative determination of neutralizing antibodies against SARS-CoV-2 variants, traceable to WHO International Standards. The test utilizes a novel Fusion Protein that mimics the Spike receptor binding domain docked to the human ACE2 protein and effectively blocks non-neutralizing antibodies in the sample. After pre-blocking sequesters the non-neutralizing antibodies from the samples, direct binding of the residual neutralizing antibodies to variant RBDs coated in the wells of the microtiter plate is measured with a fluorescent secondary antibody. Results of the Q-NAb IgG Test agree with a live virus Microneutralization Assay for both the Ancestral strain (WA1-2020) and the Omicron BA.5 (COR-22-063113/2022) variant (Spearman’s correlation, ρ = 0.87 and 0.92, respectively). The analytical performance (LoB, LoD, LoQ, linearity, precision, and interference) of the Q-NAb IgG Test was established along with sensitivity and specificity using a panel of monoclonal neutralizing and non-neutralizing anti-SARS-CoV-2 antibodies. Clinical sensitivity and specificity using pre-pandemic, convalescent, and vaccinated serum and plasma samples is also reported. The advantages of the Q-NAb IgG Test are its strong correlation to live virus neutralization tests, traceability to WHO International Standards, convenient microtiter plate format, low sample volume requirements, and suitability for a BSL-2 laboratory.

## Introduction

Multiple studies involving vaccinated and convalescent subjects have demonstrated that the titer for neutralizing antibody (NAb) is correlated with protection against COVID-19^1–3^. Recent studies with half-life extended monoclonal antibody therapy provide strong evidence that NAbs are mechanistic in mediating this protection against symptomatic COVID-19^4^. In fact, recent updates to Emergency Use Authorization of SARS-CoV-2 vaccine boosters accept NAb titer as a correlate for protection without the need for additional randomized trials to demonstrate clinical efficacy^5^.

Virus neutralization assays have largely been used to report NAb levels in vaccine trials. However, these assays are labor-intensive, time-consuming, have poor reproducibility, and suffer from a lack of standardization. Moreover, the use of live SARS-CoV-2 virus in these assays requires that they be performed in a BSL-3 laboratory. Although the World Health Organization (WHO) has released international standards for measuring NAb levels, the lack of standardized neutralization tests in all the varied studies has led to normalization of the 50% inhibition titers against study-specific convalescent patient cohorts, permitting only a relative measure of NAb titers between studies^1,6^. On the other hand, commercial surrogate virus neutralization kits that measure NAb titer by measuring the competitive inhibition of RBD-ACE2 binding by NAb offer the advantages of high-throughput sample processing and are desirable for immune surveillance and vaccine efficacy studies^7,8^. However, these assays suffer from limited assay range and, in most cases, give only a qualitative (or semi-quantitative) result^9^. Lastly, the commercial neutralization assays validated to date are designed to measure NAb against the ancestral SARS-CoV-2 strain present at the start of the global pandemic^10^. Measuring NAb titers against Omicron and other later variants requires a new strain-specific neutralization assay that is not currently available.

In this paper, we describe the design and validation of the Q-NAb IgG Test, a cell-free, direct-bind plate assay to quantify NAbs against ancestral SARS-CoV-2 and is easily adapted to measure NAb titer against other variants of concern in clinical samples. The Q-NAb IgG Test uses serum or plasma to report out a quantitative result of NAb titer (IU/mL) that is traceable to WHO standards and can be performed in a standard BSL-2 clinical lab environment. To achieve this, we developed a novel Fusion Protein (FP) that mimics the Spike receptor binding domain (RBD) docked to the human ACE2 protein, the functional receptor on cells for viral entry. The docked format of the FP is held in the closed configuration such that the receptor binding motif (RBM) is hidden due to the interaction of RBD with human ACE2 while the rest of the RBD is exposed. When incubated with a clinical sample, the FP acts as an effective blocker of non-neutralizing antibodies (NNAb). In conjunction with solid-phase bound RBD, this allows for the direct detection of NAbs in the clinical specimen. Here we report the analytical and clinical performance of two versions of the Q-NAb IgG Kit developed to detect NAb against Ancestral and BA.4/5 (the RBD sequence is identical in both BA.4 and BA.5 SARS-CoV-2 variants).

## Methods

### Protein Constructs

For transient protein expression in mammalian cell suspension culture, the RBD and RBD/ACE2 FP open reading frame constructs were each inserted into a mammalian expression vector, pcDNA3.4 (LifeScience Market). Constructs were designed with Kozak sequence preceding the start codon, for ensuring the correct translation initiation, followed by a signal peptide (mouse Ig heavy chain variable region) for protein secretion^11^. Immediately after the signal peptide, DNA encoding RBD or FP was inserted. The sequences for proteins were based on GenBank MN908947.3 for RBD and NCBI Gene ID 59272 for human ACE2. For RBD BA.4/5 and FP constructs, the C-terminus of RBD was fused with amino acid 18-118 and 18-614 of human ACE2, respectively, by a linker sequence that consists of Avi-tag, Tobacco Etch Virus (TEV) protease site, and (G4S)2. At the C-terminus of ACE2, a 6xHistidine tag was added to aid in Ni-NTA affinity purification. All of the gene sequences were synthesized by IDT using human codon biases for amino acids encoded by multiple triplets in the DNA code.

### Expression of Proteins

Plasmid encoding RBDs and FP were transfected into Expi293 cells (Thermo Fisher Scientific) by polyethyleneimine, cultured in Expi293 expression medium (Thermo Fisher Scientific) at 37°C with 5% CO_2_ for 120 hours. The supernatant containing expressed proteins were purified with Ni-NTA through His-tag binding, followed by a Superdex 75 or 200 gel filtration column (Cytiva) in 25 mM HEPES, 150 mM sodium chloride, and 5% glycerol.

### Biotinylation and Streptavidin Gel-shift Assay

After purification, in-house biotinylation of RBD proteins was performed using BirA enzyme (Avidity) at 30°C for 40 min. For BA.4/5 RBD, the biotinylated protein was cleaved using TEV protease (Sigma-Aldrich) to remove the ACE2 fragment of the construct. Free biotin was removed using a Superdex 75 column. Biotinylation efficiency was confirmed by a streptavidin gel-shift assay^12^. Briefly, biotinylated RBD proteins were mixed with a 3-fold molar excess of streptavidin and incubated for 5 minutes. The samples were then run on SDS-PAGE gel and stained with Coomassie blue. The degree of biotinylation was quantified by densitometry using ImageJ.

### ACE2 Binding Assay

To evaluate the functionality of RBD proteins, an ACE2 binding assay using size exclusion chromatographic peak analysis was conducted. Each target protein was mixed with a molar excess of ACE2 protein (2- to 3-fold) for 5 minutes. The mixture was then injected onto a Superdex 75 gel-filtration column (Cytiva) using FPLC (AKTA, GE). The peaks detected by UV absorbance at 280 nm were compared to those obtained from injections of ACE2 or target protein alone.

### Monoclonal Ab ELISA

A flat-bottom 96-well clear high-binding polystyrene microtiter plate (Corning Cat# 9018) was coated overnight at 4°C with 300 ng/mL Neutravidin (Sigma-Aldrich) and blocked using assay buffer containing 2.5% BSA (Sigma-Aldrich) in PBST (150 mM NaCl, 3 mM KCl, 8 mM Na_2_HPO_4_, 2 mM KH_2_PO_4_, and 0.05% Tween 20) for 1 h at room temperature. After washing with PBST, a 20 nM solution of biotinylated variant RBD was added to the neutravidin coated wells and incubated at room temperature for 1 hour. Commercially available biotinylated RBD proteins (AcroBiosystems) specific to the Ancestral (Cat# SPD-C82E9) and BA.4/5 (Cat# SPD-C82Ew) strains were used as positive controls. Since the FP is not biotinylated, it was directly bound to the microtiter plate. The unbound protein was removed and washed 3 times by PBST. Neutralizing and non-neutralizing commercial monoclonal antibodies were then added to each well at a concentration of 30 ng/mL in assay buffer and incubated at room temperature for 1 h. The antibody solution was removed, and the wells washed 3 times by PBST, followed by addition of HRP-conjugated anti-human IgG antibody (Sigma) at 100 ng/mL in assay buffer. After incubation at room temperature for 1 hour, the wells were washed with PBST three times followed by addition of SIGMAFAST_™_ OPD *o*-phenylenediamine dihydrochloride substrate solution (Sigma). After incubation at room temperature for 15 minutes, the OD_450_ absorbance was measured using SpectraMax iD3 microplate reader (Molecular Devices).

### Clinical Samples

Donor de-identified human serum and plasma samples were sourced from New York Biologics (Southampton, NY) and Innovative Research (Novi, MI). These samples were collected either at FDA-approved collection sites or under IRB approved protocol from participants who provided written informed consent for blood specimen collection and subsequent analysis (WCG IRB tracking number 20010304).The samples used in the validation of the Q-NAb IgG Ancestral Kit included 137 samples from individuals that received the initial mRNA vaccine series (Comirnaty or Spikevax), 51 samples from individuals that had recovered from SARS-CoV-2 infection, and 19 samples collected prior to Dec 2019 (pre-pandemic). Samples used in the validation of the Q-NAb IgG BA.4/5 Kit included 151 samples from individuals that received a bivalent vaccine booster (Comirnaty or Spikevax), 20 samples from individuals that had recovered from SARS-CoV-2 infection, and 29 samples collected prior to Dec 2019.

### Microneutralization Assay

Live-virus microneutralization assays were performed to determine the half-maximal microneutralization concentration (MN50) of clinical samples by Microbiologics (St. Cloud, MN). Vero E6 and Vero E6 TMPRSS2-T2A-ACE2 (VTA) mammalian cell lines were used to determine the MN50 against Ancestral SARS-CoV-2 (SARS-CoV2/human/USA/USA-WA1/2020) and SARS-CoV-2 BA.5 variant (hCoV-19/USA/COR-22-063113/2022), respectively.

Briefly, the clinical samples were serial diluted in viral growth medium (Dulbecco’s Modified Eagle’s Medium (DMEM) with 2% FBS supplemented with antibiotic-antimycotic, 1X Gibco MEM nonessential amino acids (MEM-NEAA), and 1 mM sodium pyruvate) and each diluted sample was mixed with 200 TCID50 of the appropriate SARS-CoV-2 strain for 1 h at 37°C and 5% CO_2_. Following the incubation, the virus-serum mixture was added to a confluent monolayer of mammalian cell culture seeded in 96-well tissue culture treated plates and incubated at 37°C in a 5% CO_2_ incubator for 2 and 7 days for Ancestral and BA.4/5 variant, respectively. Appropriate virus and cell controls were used as quality control checks.

For Ancestral strain, at 2 days post-incubation, the supernatant was removed, and the cells were washed and fixed. Primary antibodies against SARS-CoV-2 nucleoprotein were added to each well of the plate and after washing, labeled using an HRP-conjugated secondary. The plate was washed, colorimetric HRP substrate was added, and absorbance was measured at 405 nm after the addition of stop solution.

For the BA.5 variant MNA, at 7 days post-infection the cytopathic effect was observed using an inverted microscope. Cells were also stained with a freshly prepared 1:10 dilution of cell viability reagent (AlamarBlue HS, Thermo Fisher Scientific). Plates were subsequently incubated in the dark at 37°C in a 5% CO_2_ incubator for at least 4 h. A 100 µL aliquot of supernatant was transferred to a new clear bottom 96-well plate and fluorescence intensity was measured at 560 nm excitation and 590 nm emission wavelength.

### Q-NAb IgG Kit

#### Q-NAb Plate

Black 96-well Maxisorp plate (Thermo Fisher Scientific) was coated overnight with 100 µL of 6 µg/mL Neutravidin in PBS at 4°C followed by incubation with 100 µL of 200 nM of biotinylated RBD in assay buffer for 1 h at room temperature. The wells were subsequently washed thrice with 1X PBST to remove excess RBD and were blocked and stabilized with Stabilguard (Surmodics) for 1 h at room temperature. Excess Stabilguard was aspirated and the plates dried overnight in a nitrogen cabinet at room temperature before use.

#### Q-NAb IgG Detector

Donkey anti-human IgG (Jackson ImmunoResearch) was conjugated to Alexa Flour 594 (Thermo Fisher Scientific) according to manufacturer’s instructions using a 4 times molar excess of the dye. After conjugation, the excess dye was removed using a 40K molecular weight cutoff Zeba Spin desalting column (Thermo Fisher Scientific). The detector conjugate was then HPLC purified using a Superdex 200 Increase 10/300 GL column using an isocratic elution with 150 mM sodium phosphate buffer at pH 7 and the fractions collected were pooled and concentrated to 2 mg/mL. Approximately 144 µg of Alexa Fluor 594 conjugated donkey anti-human IgG was lyophilized in a 20 mL amber glass vial in a formulation containing 75 mM sodium phosphate, 12.5 mM HEPES, 15% trehalose, 0.1% mannitol, and stored at 4°C.

#### Q-NAb Fusion Protein

Approximately 160 µg of FP was lyophilized in a 20 mL clear glass vial in a formulation containing 25 mM HEPES, 250 mM NaCl, 15% trehalose, 0.1% mannitol, and stored at 4°C until use.

#### Q-NAb IgG Calibrator and Control

The calibrator was prepared by lyophilizing pooled high titer serum samples in a 2 mL glass vial in a formulation containing 12.5 mM HEPES pH 7.5, 15% trehalose, 0.1% mannitol, 0.3% tributyl phosphate, 1% Tween-80, and stored at 4°C. Q-NAb Positive External Control for the BA.4/5 Kit was prepared by lyophilizing 77.5 ng of anti-RBD-BA.4/5 monoclonal NAb (clone: 10B1A5, AcroBiosystems) in a formulation containing 1X PBS (10mM phosphate buffer pH 7.4, 0.15mM NaCl, 3mM KCl), 1% BSA, 10% trehalose, in a 2 mL clear glass vial. Q-NAb Positive External Control for the Ancestral Kit was prepared by lyophilizing ∼100 ng of anti-RBD NAb (clone: AM122, AcroBiosystems) in the above formulation.

### Calibrator Value Assignment and Traceability to WHO International Standard

The neutralization titer of Q-NAb IgG Calibrators was assigned based on appropriate WHO International Standards (National Institute of Biological Standards and Controls, UK). Specifically, the Q-NAb IgG Ancestral Kit is calibrated and traceable to the 2^nd^ International Standard for anti-SARS-CoV2 immunoglobulin (NIBSC code 21/340) and the Q-NAb IgG BA.4/5 Kit is calibrated and traceable to the 1^st^ International Standard for antibodies to SARS-CoV2 variants of concern (NIBSC code 21/338). Briefly, each lower-level calibrator (below the international standard in calibrator hierarchy) is value assigned from a higher-level calibrator by testing a two-fold dilution of both calibrators on the same plate. For example, a two-fold dilution series of master calibrator and WHO International Standard are tested in triplicate (N=5 for lower dose levels) on the same plate and the weighted average dose of the master calibrator was calculated from the 4-PL fit of the WHO Standard. To minimize overall uncertainty in the assigned value of the master calibrator, value assignment was repeated using three independent lots of Q-NAb IgG Kits and the average from the three lots was the assigned value of the master calibrator. A similar process was used to value assign the end-user calibrator using the master calibrator. The total uncertainty in the value assignment of the calibrators was calculated in accordance with ISO 17511:2020 standard. End-user calibrator was assayed on every plate to provide direct traceability of Q-NAb neutralization titer to the respective WHO International Standards.

### Q-NAb IgG Test

The Q-NAb Plates were washed to remove excess blocker/stabilizer before use. Lyophilized vials of FP and detector were reconstituted in 20 mL and 12 mL of assay buffer, respectively. Samples tested using the Q-NAb IgG BA.4/5 Kit were diluted 1:20 in reconstituted FP for 10 – 15 min at room temperature to deplete NNAb. Samples tested using the Q-NAb IgG Ancestral Kit were similarly treated using a starting dilution of 1:25. Lyophilized calibrator was reconstituted using 620 µL of reconstituted FP (775 µL for Ancestral Kit) and a 1:2.5 dilution series was prepared. Lyophilized external control was also similarly reconstituted. Both calibrator and control were incubated for 10 – 15 min at room temperature taking care to ensure that they were incubated for the same time as the diluted samples. All depleted samples, calibrator and controls were tested in triplicate by adding 100 µL/well and incubating shaking at room temperature for 1 h. The plate was washed thrice with 1X PBST and incubated with 100 µL/well of reconstituted detector for 1 h shaking at room temperature. After a final wash to remove the excess detector, fluorescence was measured using 560 nm excitation and 620 nm emission wavelengths. Raw fluorescence signal for samples and controls were converted to NAb titer using a 4-PL fit of the plate-specific calibration curve using previously assigned calibrator values for the respective assays (BA.4/5 and Ancestral). Pre-specified plate acceptance criteria and control dose recoveries were applied as part of robust quality control.

### Analytical Performance of Q-NAb IgG Kits

The analytical performance characteristics were established in accordance with applicable CLSI Standards for both the Ancestral and BA.4/5 Q-NAb IgG Kits. Briefly, the following performance metrics were evaluated for each kit:

a. Limit of Blank (LoB): Five pre-pandemic serum samples were each tested in triplicate on two days using two lots of Q-NAb IgG Kits. The limit of blank for each lot was evaluated as the 95^th^ percentile of the 30 total measurements. The larger of the two limits of blank is reported as the Limit of Blank of the Q-NAb IgG Kit.
b. Limit of Detection (LoD): Five low-level positive samples were each tested in triplicate on each of two days using two lots of Q-NAb IgG Kits. The limit of detection for each kit was determined in accordance with CLSI EP17-A2. The larger of the two is reported as the Limit of Detection of the Q-NAb IgG Kit.
c. Limit of Quantitation (LoQ): Twenty replicates of each of four low-level samples were assayed on each of two lots of Q-NAb IgG Kits. In accordance with CLSI EP17-A2, for each lot, the lowest dose where the CV is less than 20% was determined as the LoQ of that lot. The larger of the two LoQ is reported as the LoQ of the Q-NAb IgG Kit.
d. Linearity: A dilution series from a high sample pool was prepared in assay buffer and tested in triplicate. In accordance with CLSI-EP06-Ed2, a weighted least square regression was used to evaluate the linearity of the Q-NAb IgG Kit.
e. Precision: Five samples (4 clinical samples/sample pools and one external control) across the analytical range of the assay were tested in a 20x2x2 single-site precision study. In accordance with CLSI EP05-A3, a single lot of Q-NAb IgG Kit was used to evaluate repeatability, between-day, and intermediate precision of the assay.
f. Interference Screening: A serum sample with a neutralizing titer of ∼7X LoQ of the Q-NAb IgG Kit was used to screen for potential interference from endogenous and common exogenous substances. Briefly, the sample was spiked with potential interfering substance from a 20X stock of the interferent (in appropriate diluent). This serum sample spiked with the potential interference and a corresponding sham spike (spiked with only diluent) were tested in replicates of N=10 to detect a 15% difference in dose recovery between the interferent and sham-spiked samples.

### Statistical analysis

A commercial 4-PL utility tool (Graphpad Software) was used to convert raw fluorescence signal to Q-NAb IgG neutralization titer. The correlation of MN50 values and Q-NAb IgG neutralization titer was analyzed using JMP statistical software suite. Analytical performance characteristics of Q-NAb IgG Kit were analyzed using method validation add-in for MS Excel (Analyse-It).

## Results

### Design and Expression of RBD and FP

The defining feature of the Q-NAb technology is the design of the RBD-ACE2 FP. The FP is made of the SARS-CoV-2 Spike RBD docked to the human ACE2 protein, the functional receptor on cells for viral entry. The FP construct encodes the BA.4/5 RBD at the N-terminus with the ACE2 at the C-terminal end of a single polypeptide (Figure S1). To allow the unconstrained intramolecular binding of the RBM to theACE2, a flexible amino acid linker was also encoded between the C-terminal end of the RBD and the N-terminal end of the ACE2. To further stabilize the RBM/ACE2 interaction and to hold the FP in a closed configuration, a disulfide bond was engineered into the interface to create a disulfide bridge. Specifically, AA38 of the ACE2 (Glu) and AA496 on the RBD (Gly) when mutated to cysteines minimized the distance (1.7 Å) between the cysteine side chains without steric clash (91degree dihedral angle). In the closed configuration, the Receptor Binding Motif (RBM) is kept hidden (inaccessible) due to its stable interaction with ACE2 while the rest of the RBD is exposed.

The ancillary piece of the Q-NAb technology is the design of the ancestral and variant RBDs (Figure S2 and S3). The recombinant RBD polypeptide was engineered to have a conserved region to code the region away from the RBM (shown in blue in Figure 1) and a variant RBM region (shown in red in Figure 1) that carried all the consensus mutations found in the prevalent circulating strain of the SARS-CoV-2 variant. Table 3 lists all the consensus mutations incorporated into recombinant variant RBDs produced using the Q-NAb technology. Using a conserved sequence for the non-RBM region of the RBD enabled us to not only produce these variant RBDs at a high yield but also enabled the Fusion Protein (using the same conserved non-RBM region) to be used as a universal blocker for NNAb.

**Figure 1:**
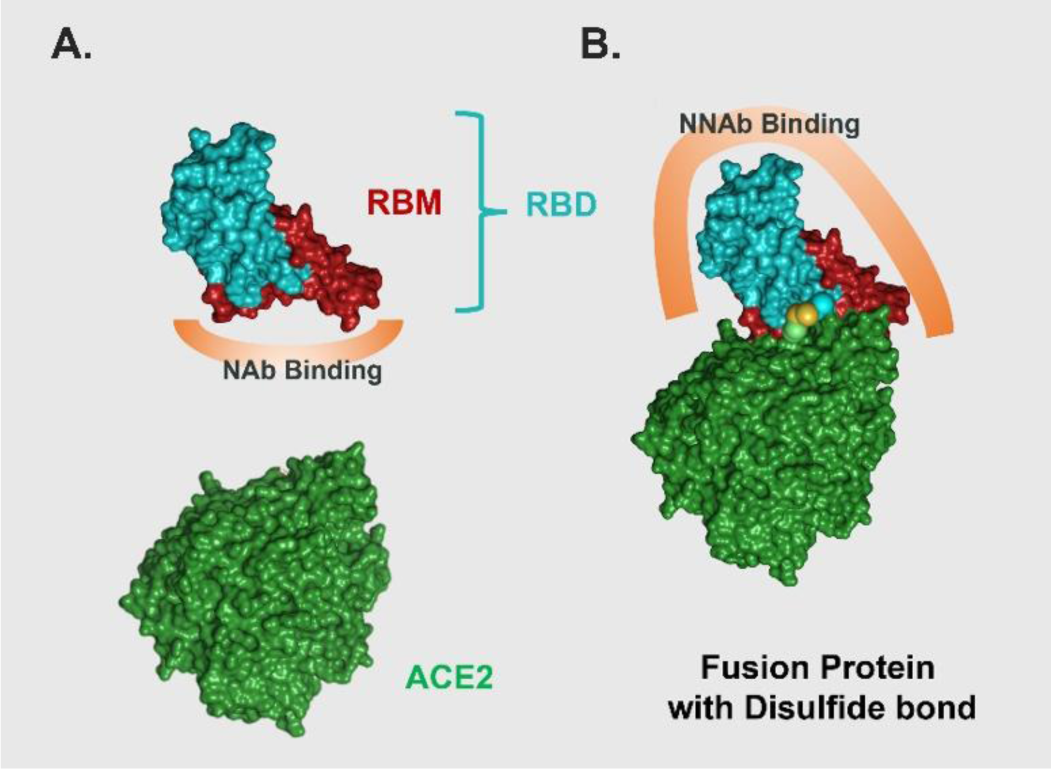
Undocked (A) and docked (B) structures of RBD-ACE2 FP. Different domains are color coded; the engineered disulfide linkage is shown in yellow and the putative NAb and NNAb binding regions are identified.

**Table 1:**
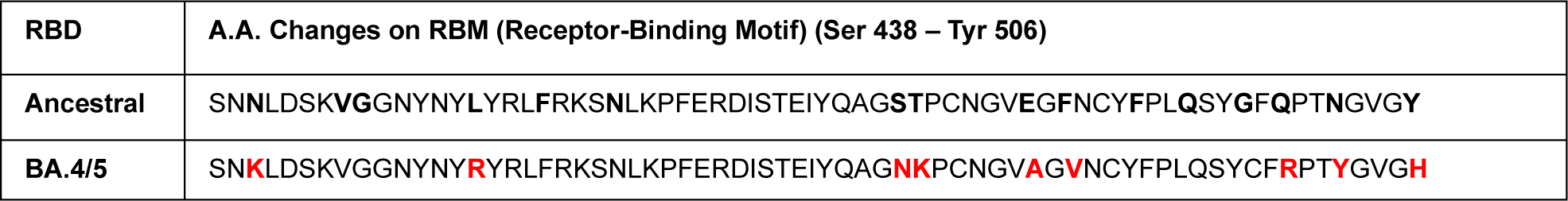
Amino acid sequence showing the consensus sequence (mutations in BA.4/5 are shown in red) in the RBM region of SARS-CoV-2 (Ancestral and BA.4/5) engineered into a backbone of conserved non-RBM RBD residues to enable high yield of recombinant RBDs in transient HEK293 mammalian cell culture.

### Protein Functionality

The functionality of recombinantly produced RBD (Ancestral and BA.4/5) and FP were tested using 14 different commercially available variant-specific NAbs and NNAbs using an end-point in-house assay (Table 2). As expected, all three proteins bound the NNAbs that were tested to a similar degree (Figure S2 and S3). In contrast, there were expected differences in the binding profile of the proteins to NAbs. The FP did not bind any of the NAbs. This is expected because, in the closed configuration, the receptor binding motif of the RBD is shielded by ACE-2 portion of the FP. The recombinant ancestral RBD bound a wide-range of commercial NAbs that recognizes binding epitopes in the Ancestral RBD. The recombinant BA.4/5 RBD bound only one commercially available BA.4/5 specific NAb. Taken together, these binding profiles show that the FP could be used selectively to block and deplete the NNAbs without affecting/depleting the NAbs in the sample. In addition, the availability of the commercial NNAbs and NAbs provided a straight-forward quality control tool for production of the recombinant proteins in-house.

**Table 2:** Binding of Recombinant RBD and FPs to commercially available NNAb and variant-specific NAb.

| Antibody Type | Vendor/Source | RBD Ancestral | RBD BA.4/5 | Fusion BA.4/5 |
| --- | --- | --- | --- | --- |
| **NNAb** | Genescript HC2001 | + | + | + |
|  | Hytest RBD5305 | + | + | + |
|  | InVivogen CR3022 | + | + | + |
|  | ActiveMotif AM015553 | + | + | + |
|  | ActiveMotif AM006415 | + | + | + |
|  | ActiveMotif AM038105 | + | + | + |
|  | AcroBiosystem AM130 | + | + | + |
| **NAb** | AcroBiosystems AM122 | + | - | - |
|  | ActiveMotif AM009105 | + | - | - |
|  | ActiveMotif AM005505 | + | - | - |
|  | ActiveMotif AM001414 | + | - | - |
|  | InVivoGen Casirivimab-derived | + | - | - |
|  | InVivoGen Imdevimab-derived | + | - | - |
|  | AcroBiosystems 10B1A5, BA.4/5 Specific | - | + | - |

### Q-NAb IgG Test

The Q-NAb IgG Test measures the NAb titer in a sample by first sequestering the NNAbs using the FP and enriching the NAbs in the sample. Variant-specific NAbs are then detected using a direct assay where the wells are coated with the variant RBD and yield a signal that is directly proportional to the concentration of NAb. The quantitative Q-NAb IgG Kit consists of a microtiter plate coated with variant specific RBD protein and lyophilized vials of FP, calibrator, positive external control, and fluorescent conjugated detector.

**Figure 2:**
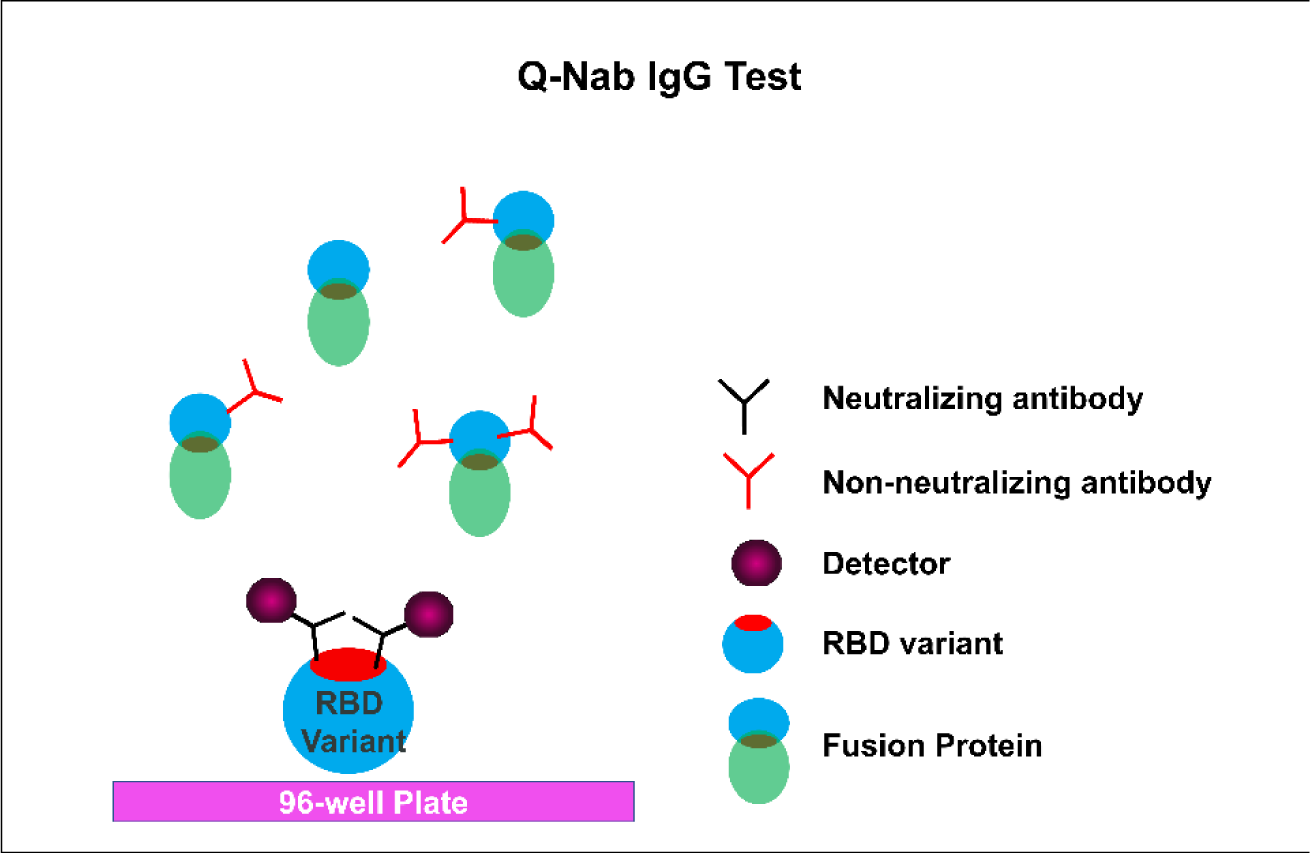
The quantitative Q-NAb IgG Test was designed to directly detect NAbs binding to SARS-CoV-2 RBD variants on the microplate after sequestering NNAbs using FP.

### Validation of the Ancestral and BA.4/5 Q-NAb IgG Kits

To validate the Q-NAb IgG Test platform, we compared the Q-NAb neutralization titer against gold standard CPE-based microneutralization assay (MNA) using a wide range of clinical specimens including vaccinated, convalescent, and pre-pandemic samples (Table 3). As a qualitative assay, both Q-NAb assays (Ancestral and BA.4/5) had a >94% sensitivity while the specificity of the Q-NAb Ancestral and BA.4/5 assays were 91% and 54%, respectively. The low specificity of the Q-NAb BA.4/5 assay was exclusively due to the detection of low-level BA.4/5 neutralizing activity (∼2X LoQ) of pre-omicron convalescent samples that was not detected using the MNA (Table 3). In fact, the pre-omicron convalescent samples had a median BA.4/5 neutralization titer of 750 IU/mL, almost 10-fold lower than the median titer of ∼8600 IU/mL observed in samples from patients receiving a bivalent booster. As a quantitative assay, we observed a strong correlation when Q-NAb neutralization titer was compared against the MN50 values for the BA.4/5 and Ancestral assays with a Spearman’s correlation (ρ) of 0.92 and 0.87, respectively (Figure 3).

**Table 3:**
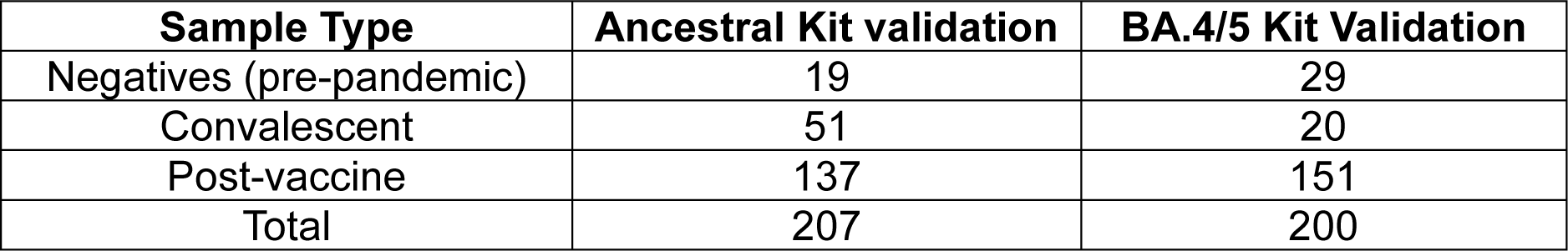
Clinical sample sets used for validation of Q-NAb IgG Kits.

**Figure 3.**
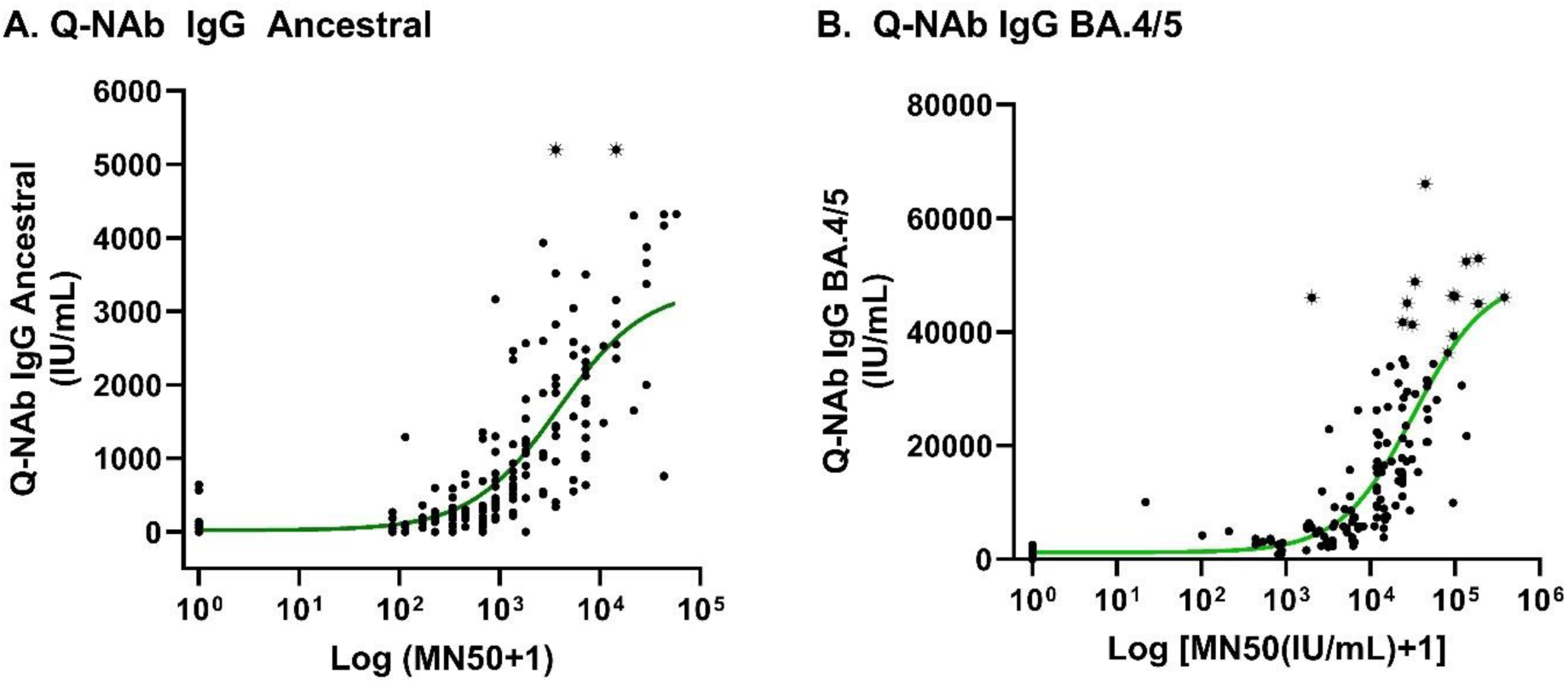
Scatter plot and parametric curve fit between Q-NAb IgG Ancestral (A) and BA.4/5 (B) neutralization titer and corresponding microneutralization assay MN50 values of clinical positive and pre-pandemic samples.

We evaluated the effect of seven potentially interfering endogenous substances and three commonly used drugs on the Q-NAb IgG Tests and found that the Q-NAb neutralization titer was not affected by either lipemic, icteric or hemolyzed samples or commonly used analgesics that is routinely used to manage COVID-19 symptoms (Table S1).

### Analytical Performance of Q-NAb IgG Kits Calibration and Traceability

The dynamic ranges for Q-NAb IgG Ancestral and BA.4/5 Kits were established to be 64 - 5500 IU/mL and 300-32000 IU/mL, respectively. Two separate WHO International Standards have been established for the calibration and harmonization of serological assays detecting anti-SARS-CoV-2 neutralizing antibodies viz. NIBSC 21/340 for early isolates and NIBSC 21/338 for variants of concern (VOCs) circulating during or after 2022. Our in-house calibrator, prepared by pooling serum from 10 individuals that were screened for high NAb titers, were value assigned using the calibration hierarchy shown in Figure 4. Table 4 shows the assigned value of the master and end-user calibrators for both Q-NAb IgG Kits and the associated cumulative uncertainties.

**Figure 4:**
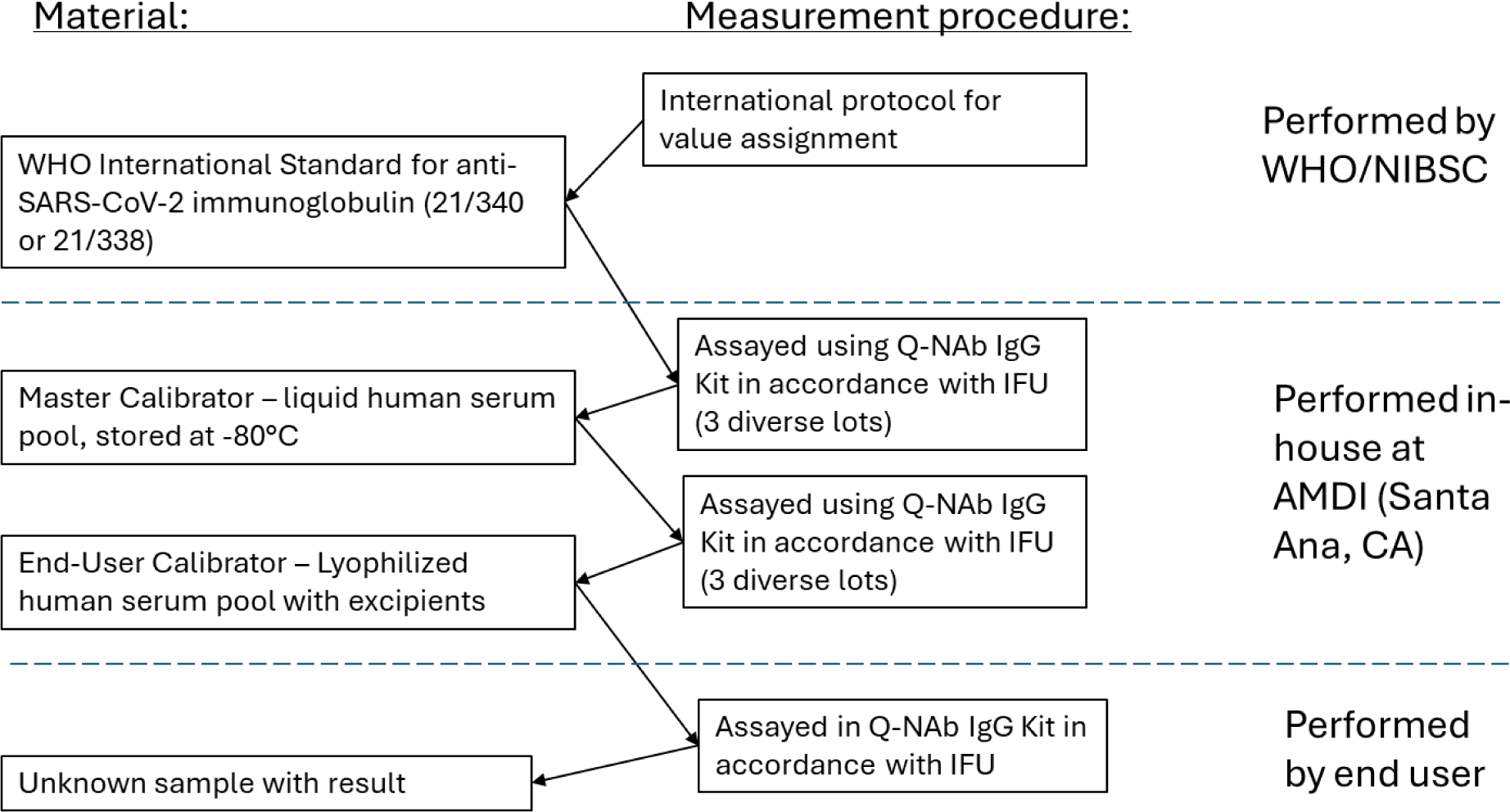
Calibration hierarchy to establish traceability of in-house master and end-user calibrators to WHO International Standard 21/340 and 21/338 for the Ancestral and BA.4/5 Q-NAb IgG Kits, respectively.

**Table 4:** Assigned value and total uncertainty of in-house master and end-user calibrators of Q-NAb IgG Kits.

|  | Ancestral Calibrator |  | BA.4/5 Calibrator |  |
| --- | --- | --- | --- | --- |
|  | Assigned Value (IU/mL) | Total Uncertainty (%) | Assigned Value (IU/mL) | Total Uncertainty (%) |
| In-house Master Calibrator | 5492 | 5.7 | 35900 | 4.3 |
| End-User Calibrator | 4578 | 8.2 | 31663 | 5.3 |

### Low-End Sensitivity

The limit of blank, limit of detection, and limit of quantitation were determined using a panel of low-level and pre-pandemic clinical samples. The analytical sensitivity of the Q-NAb IgG Kits is summarized in Table ***5***.

**Table 5:** Low end sensitivity of Q-NAb IgG Kits.

| Sensitivity | Concentration (IU/mL) |  |
| --- | --- | --- |
|  | Ancestral | BA.4/5 |
| Limit of Blank | 30 | 132 |
| Limit of Detection | 42 | 170 |
| Limit of Quantitation | 64 | 293 |

### Precision

Assay repeatability and total precision were determined by repeat testing of 5 samples (4 clinical and 1 external control) spanning the analytical measurement interval of each Q-NAb IgG Test over 20 days. Repeatability for both Q-NAb IgG Ancestral and BA.4/5 assays ranged from 3 – 8% and the intermediate precision (including between run/day components) ranged between 5 – 11% (Table 6 and Table 7).

**Table 6:** Repeatability and Intermediate precision of Q-NAb IgG Ancestral Kit.

| Samples | Mean (IU/mL) | Repeatability (%) | Intermediate precision* (%) |
| --- | --- | --- | --- |
| External Control | 166 | 3.3 | 5.6 |
| Human Serum 1 | 210 | 8.1 | 11.3 |
| Human Serum 2 | 929 | 2.9 | 5.1 |
| Human Serum 3 | 1388 | 2.5 | 5.2 |
| Human Serum 4 | 2598 | 3.6 | 7.6 |

**Table 7:** Repeatability and Intermediate precision of Q-NAb IgG BA.4/5 Kit.

| Samples | Mean (IU/mL) | Repeatability (%) | Intermediate precision (%) |
| --- | --- | --- | --- |
| External Control | 1213 | 5.4 | 10.2 |
| Human Serum 1 | 2683 | 3.1 | 7.6 |
| Human Serum 2 | 6837 | 4.7 | 7.8 |
| Human Serum 3 | 9873 | 3.2 | 7.6 |
| Human Serum 4 | 19172 | 6.4 | 11.4 |

### Linearity

As shown in Figure 5, the results obtained with serial dilution of high-titer sample pool covering the analytical measurement interval of the Q-NAb IgG Ancestral and BA.4/5 Tests had excellent agreement between the observed and expected neutralization titers. This was evident both in the correlation coefficient (R^2^ = 0.99 for both) and the equations of the regression lines which were close to the respective lines of identity. Furthermore, the intercepts were less than the limits of blank for the respective Tests, in-line with the low-end sensitivity determined earlier.

**Figure 5:**
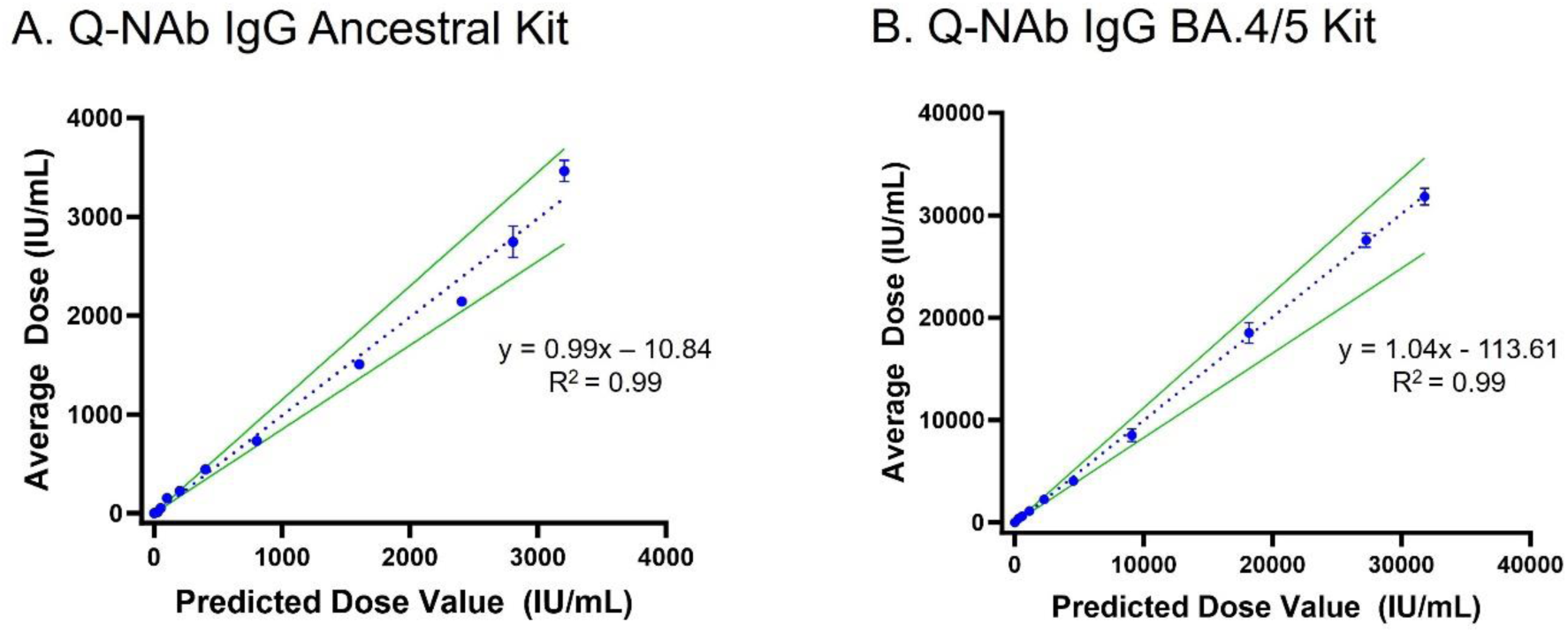
Dilution linearity of Q-NAb IgG Ancestral (A) and BA.4/5 (B) Kit using a serum pool having a high NAb titer (N=3). Weighted least square analysis was used to estimate deviation from linearity and simple linear regression analysis used to estimate fit parameters and correlation coefficient.

## Discussion

In the wake of the COVID-19 pandemic, virus neutralization levels against circulating SARS-CoV-2 strains have emerged as an important surrogate to assess population-level and individual seroprotection. This information is particularly relevant to guide future vaccination efforts in underserved communities as well as identify vulnerable individuals who are potentially immune compromised. Commercially available antibody tests measuring anti-SARS-CoV-2 IgG were developed and validated on the ancestral SARS-CoV-2 strain raising concerns over their continued use to estimate humoral response against current VOCs. Furthermore, the first-generation serological assays specifically designed to measure NAbs were developed as competitive assays and inherently have limited analytical measurement range and are not amenable to multiplexing with additional immune markers.

In this report, we have described the design and development of the Q-NAb IgG Test, a novel quantitative immunofluorescent NAb assay for high-throughput analysis of patient samples in a BSL-2 setting. In this assay, serum samples are diluted and pre-incubated with a FP made by covalently coupling SARS-CoV-2 BA.4/5 RBD and human ACE2 protein. This blocking step is designed to selectively deplete the NNAbs in the sample, thereby enriching the NAbs in the sample. For the FP to be an effective blocker of NNAbs and not interfere with NAb binding to the RBD on the solid phase, it needs to be in a stable “closed” configuration where the RBM interaction with human ACE2 is locked in place. We accomplished this by engineering a di-sulfide bridge at the interface of this interaction while the remainder of the RBD is accessible to bind NNAbs. After a brief preincubation of the sample with the FP, variant-specific NAbs are detected in a 96-well plate that are coated with the corresponding RBD and yield a signal that is directly proportional to the concentration of NAb. Since the RBM is the business end of the interaction of the Spike protein with human-ACE2, we designed our recombinant BA.4/5 RBD to have all the consensus RBM mutations on a conserved RBD backbone corresponding to the ancestral strain. This allowed us to achieve a high-yield protein manufacturing process while preserving the variant-specific binding motif crucial for measuring NAb titer.

The gold-standard microneutralization assays that have been the mainstay of vaccine approvals show a strong correlation with the Q-NAb IgG Tests (ρ = 0.87 and 0.92 for Ancestral and BA.4/5, respectively). Although there have been numerous studies measuring neutralizing titers in different patient cohorts, until recently, there has been a lack of standardization that made it challenging to do a meta-analysis of these results. The Q-NAb IgG Kits are calibrated and traceable to WHO International Standards (NIBSC 21/340 and 21/338), which allows the quantitative comparison of results across multiple studies and different labs.

The low-end sensitivity of the Q-NAb IgG Ancestral Kit was 64 IU/mL, a level sufficient to detect a NAb titer that provides greater than 90% vaccine efficacy as observed in the COVE phase 3 trial of the mRNA-1273 COVID-19 vaccine^3^. We are not aware of any studies that have established a clinically relevant cutoff for neutralizing titers after a bivalent vaccine. We believe that the low-end sensitivity of the Q-NAb IgG BA.4/5 Kit is sufficient for risk-stratification of vulnerable and high-risk population based on their NAb titer. Both assays show excellent dilutional linearity allowing for potentially expanding the dynamic range of the assay based on laboratory needs. For example, vaccine studies might require screening potential vaccine candidates based on eliciting strong humoral response and might require sample dilution to estimate neutralizing titer.

Although Q-NAb IgG platform offers many advantages over traditional PRNT assays and first generation NAb assays, the limitations of this study need to be acknowledged. Analysis from the COVE study showed that NAbs mediate about two thirds of the observed vaccine efficacy and additional immune markers will be required to fully explain the observed effect. Similarly, other immune functions that affect neutralization titer in gold-standard MN50 assays are not measured in this platform and could explain the outliers seen in the clinical correlation samples. Second, the design of the FP around a conserved consensus non-RBM sequence will need to be revisited if future variants have multiple mutations in this region and renders the FP unable to sequester the NNAbs leading to falsely elevated neutralization titers. Notwithstanding these, the Q-NAb IgG platform is a compelling alternative to traditional measures of serodiagnosis for SARS-CoV-2.

## Conclusion

Quantitative measurement of NAb levels to SARS-CoV-2 is challenging due to the cost, turnaround time, and requirement of a BSL-3 lab for accurate results. The Q-NAb IgG Kit is a variant-specific immunofluorescent assay that has <24h turnaround time and can be performed on standard microtiter plate equipment in a BSL-2 clinical laboratory. The results obtained from the Q-NAb IgG Test show strong correlation to gold-standard live virus tests and may be useful in identifying individuals who have not had an adequate response to vaccination due to a number of host or environmental factors. Future studies will address the utility of the Q-NAb IgG Test in these patient populations.

## Data Availability

All raw data produced in the present study are available upon reasonable request to the authors. All processed data in the present work are contained in the manuscript

## Notes

### Competing Interest Statement

The authors have declared no competing interest.

### Funding Statement

The study did not receive any funding

### Author Declarations

WCG IRB gave ethical approval for collection of samples used in this work

