## Supplementary material for "Development and Validation of Novel Cell-free Direct Neutralization Assay for SARS-CoV-2": Suppl Figs and Table

**Supplementary Figures and Tables**

**Figure S1. Expression and purification of RBD-ACE2 FP.** A) Amino acid sequence of RBD-ACE2 FP. The signal peptide, Avi-tag, TEV protease site, and the hexa histidine-tag are underlined. B) RBD-ACE2 FP was purified by size exclusion chromatography (SEC) using a Superdex 200 16/600 column (Cytiva). Fractions C5 – D15 of the main peak (identified by black arrow) were collected. C) The purity of RBD-ACE2 FP was determined by 12% SDS-PAGE gel after FPLC chromatography.

A. MGWSCIILFLVATATGVHSRVQPTESIVRFPNITNLCPFGEVFNATRFASVYAWNRKRISNCVADYSVLYNSASFSTFKCYGVSPTKLNDLCFTNVYADSFVIRGNEVSQIAPGQTGNIADYNYKLPDDFTGCVIAWNSNKLDSKVGGNYNYRYRLFRKSNLKPFERDISTEIYQAGNKPCNGVAGVNCYFPLQSYCFRPTYGVGHQPYRVVVLSFELLHAPATVCGPKKSTNLVKNKSVNFGGSGLNDIFEAQKIEWHEGGSENLYFQGGSGGGGSGGGGSSQSTIEEQAKTFLDKFNHEAECLFYQSSLASWNYNTNITEENVQNMNNAGDKWSAFLKEQSTLAQMYPLQEIQNLTVKLQLQALQQNGSSVLSEDKSKRLNTILNTMSTIYSTGKVCNPDNPQECLLLEPGLNEIMANSLDYNERLWAWESWRSEVGKQLRPLYEEYVVLKNEMARANHYEDYGDYWRGDYEVNGVDGYDYSRGQLIEDVEHTFEEIKPLYEHLHAYVRAKLMNAYPSYISPIGCLPAHLLGDMWGRFWTNLYSLTVPFGQKPNIDVTDAMVDQAWDAQRIFKEAEKFFVSVGLPNMTQGFWENSMLTDPGNVQKAVCHPTAWDLGKGDFRILMCTKVTMDDFLTAHHEMGHIQYDMAYAAQPFLLRNGANEGFHEAVGEIMSLSAATPKHLKSIGLLSPDFQEDNETEINFLLKQALTIVGTLPFTYMLEKWRWMVFKGEIPKDQWMKKWWEMKREIVGVVEPVPHDETYCDPASLFHVSNDYSFIRYYTRTLYQFQFQEALCQAAKHEGPLHKCDISNSTEAGQKLFNMLRLGKSEPWTLALENVVGAKNMNVRPLLNYFEPLFTWLKDQNKNSFVGWSTDWSPYAGGHHHHHH

B.

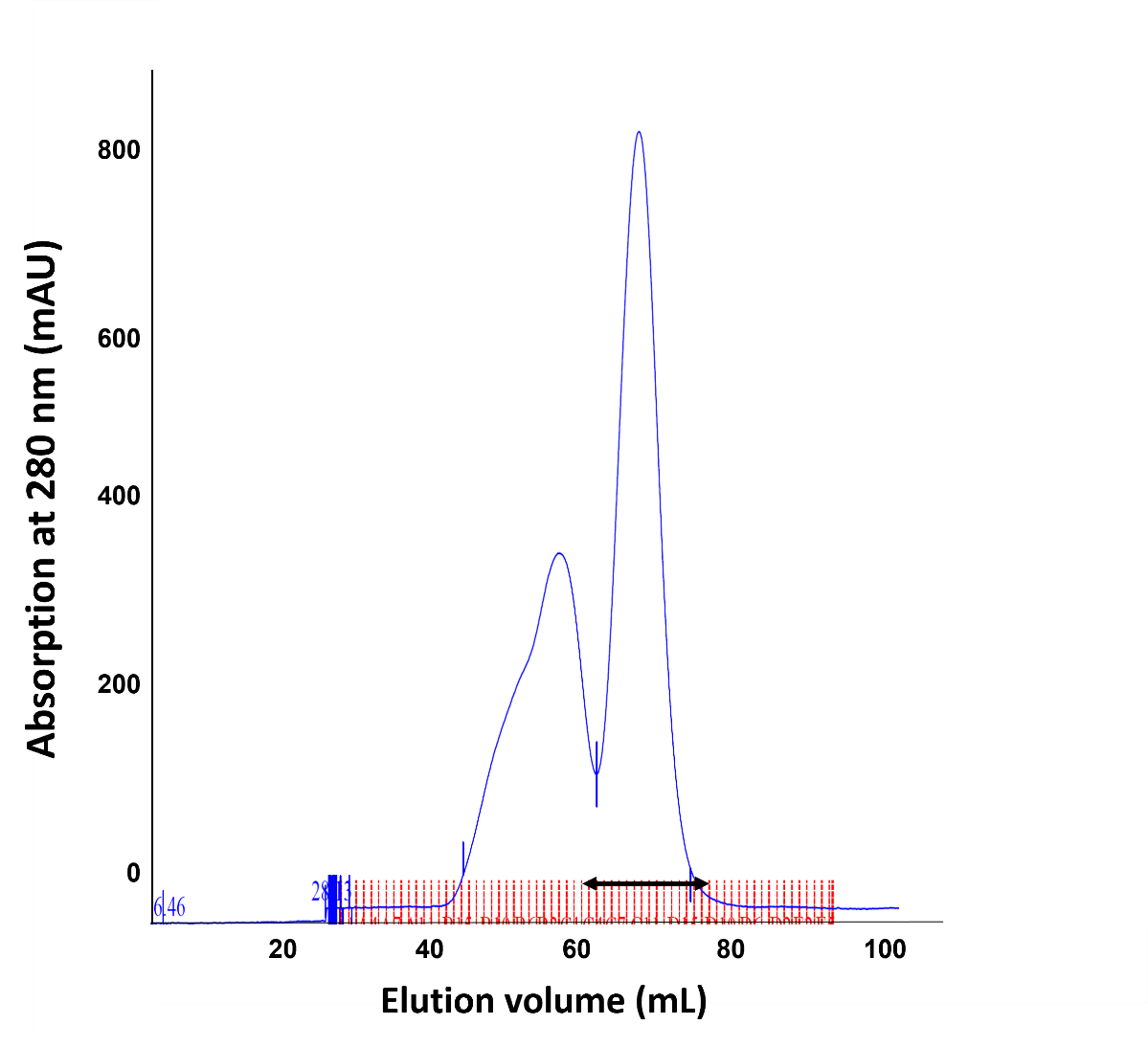

C.

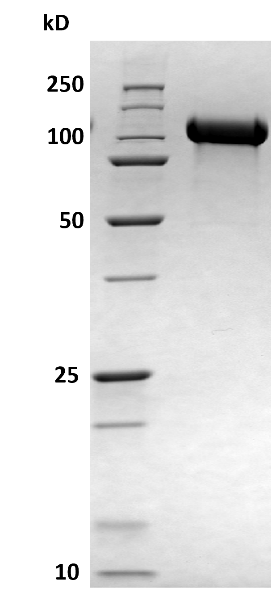

**Figure S2. Expression and purification of biotinylated SARS-CoV-2 Ancestral RBD protein.** A) Amino acid sequence of the Ancestral RBD protein. The signal peptide and the hexa histidine-tag (6XHis) are underlined. B) Streptavidin gel shift assay was done in 12% SDS-PAGE gel and 98% of protein was biotinylated based on densitometry (ImageJ) analysis. Lane 1, Ancestral RBD only (2.5 µM). Lane 2, Ancestral RBD (2.5 µM) with Streptavidin (8.3 µM). Lane 3, Streptavidin only (8.3 µM). C) Ancestral RBD was purified by size exclusion chromatography (SEC) using a Superdex 75 16/600 column (Cytiva). Fractions B7 – C5 of the main peak (identified by black arrow) were collected. D) The purity of Ancestral RBD protein was determined by 12% SDS-PAGE gel after FPLC chromatography. E) The ACE2 binding of Ancestral RBD was evaluated by adding molar excess of ACE2 to RBD and analyzing the mixture by SEC using Superdex 200 Increase 5/150 column (Cytiva). By comparing the peak areas of RBD (~2.1 mL elution volume), we found that 93% of Ancestral RBD was bound to ACE2.

MGWSCIILFLVATATGVHSRVQPTESIVRFPNITNLCPFGEVFNATRFASVYAWNRKRISNCVADYSVLYNSASFSTFKCYGVSPTKLNDLCFTNVYADSFVIRGDEVRQIAPGQTGKIADYNYKLPDDFTGCVIAWNSNNLDSKVGGNYNYLYRLFRKSNLKPFERDISTEIYQAGSTPCNGVEGFNCYFPLQSYGFQPTNGVGYQPYRVVVLSFELLHAPATVCGPKKSTNLVKNKCVNFHHHHHH

B.

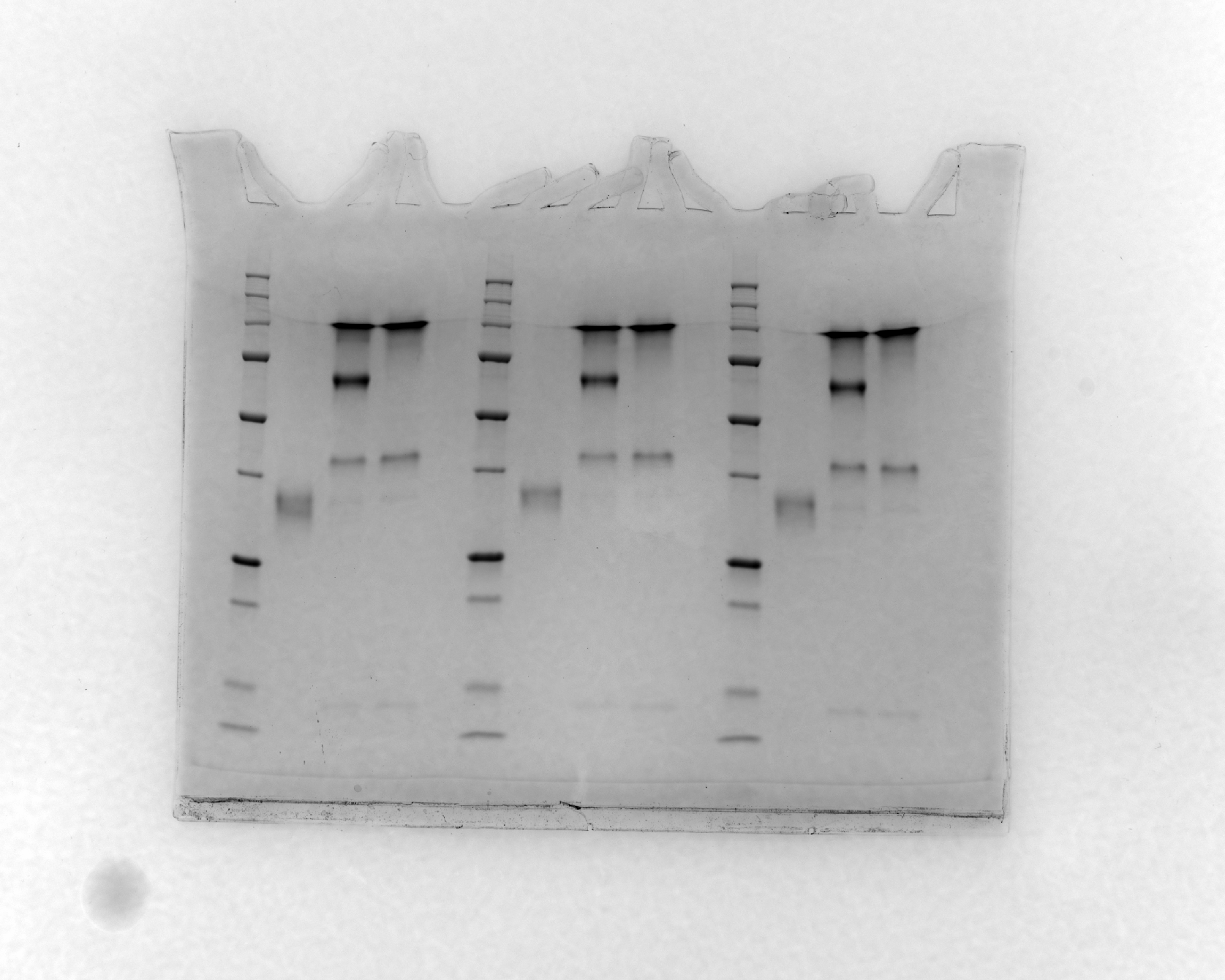

**1	2	3**

**Streptavidin bound biotinylated RBD**

**Biotinylated RBD**

**25**

**37**

**50**

**75**

**100**

**kD**

C.

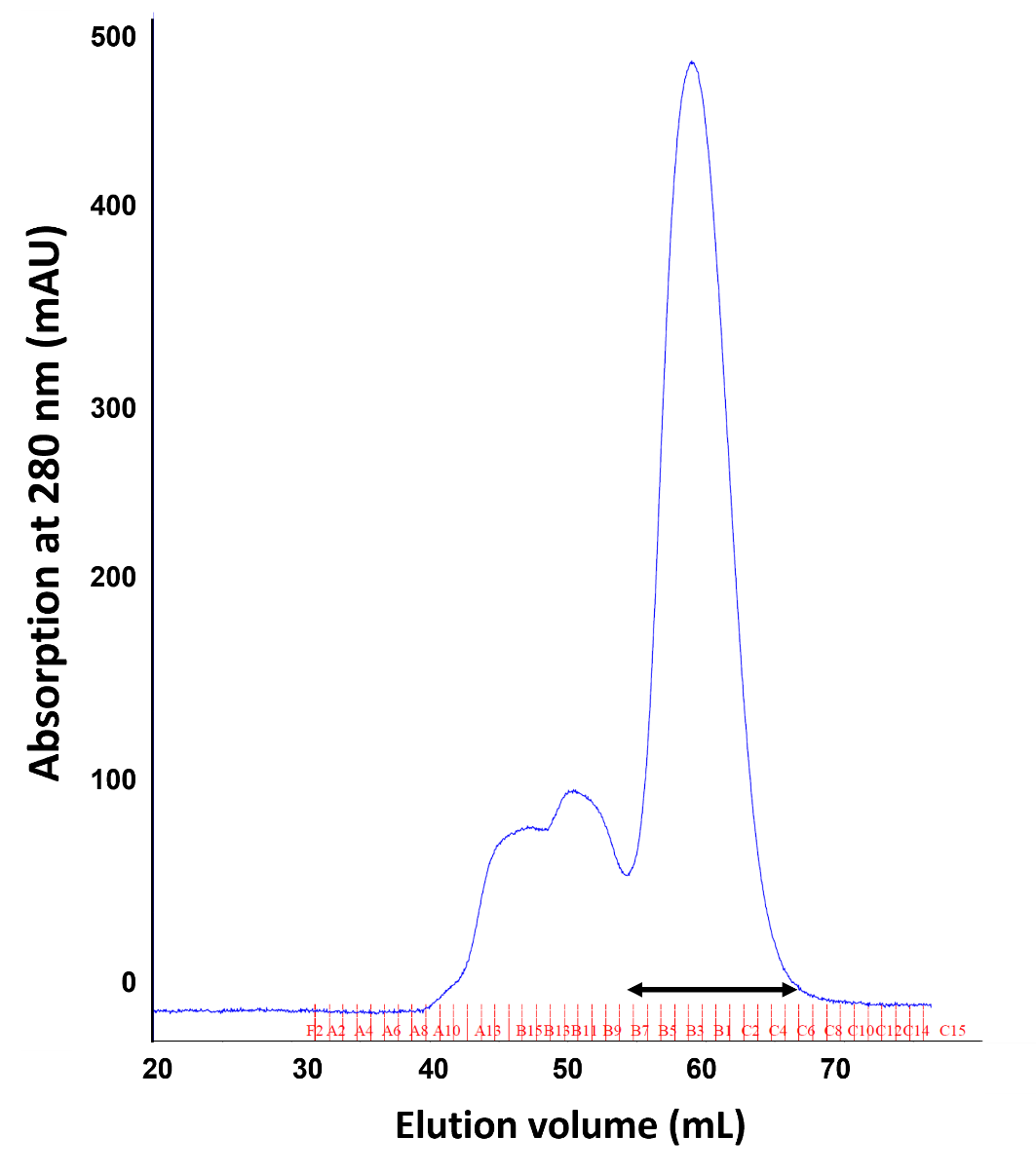

D.

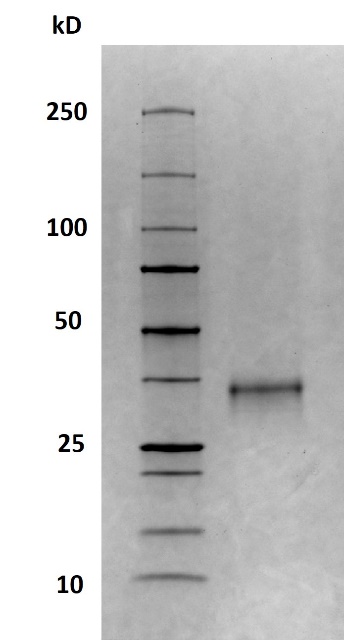

E.

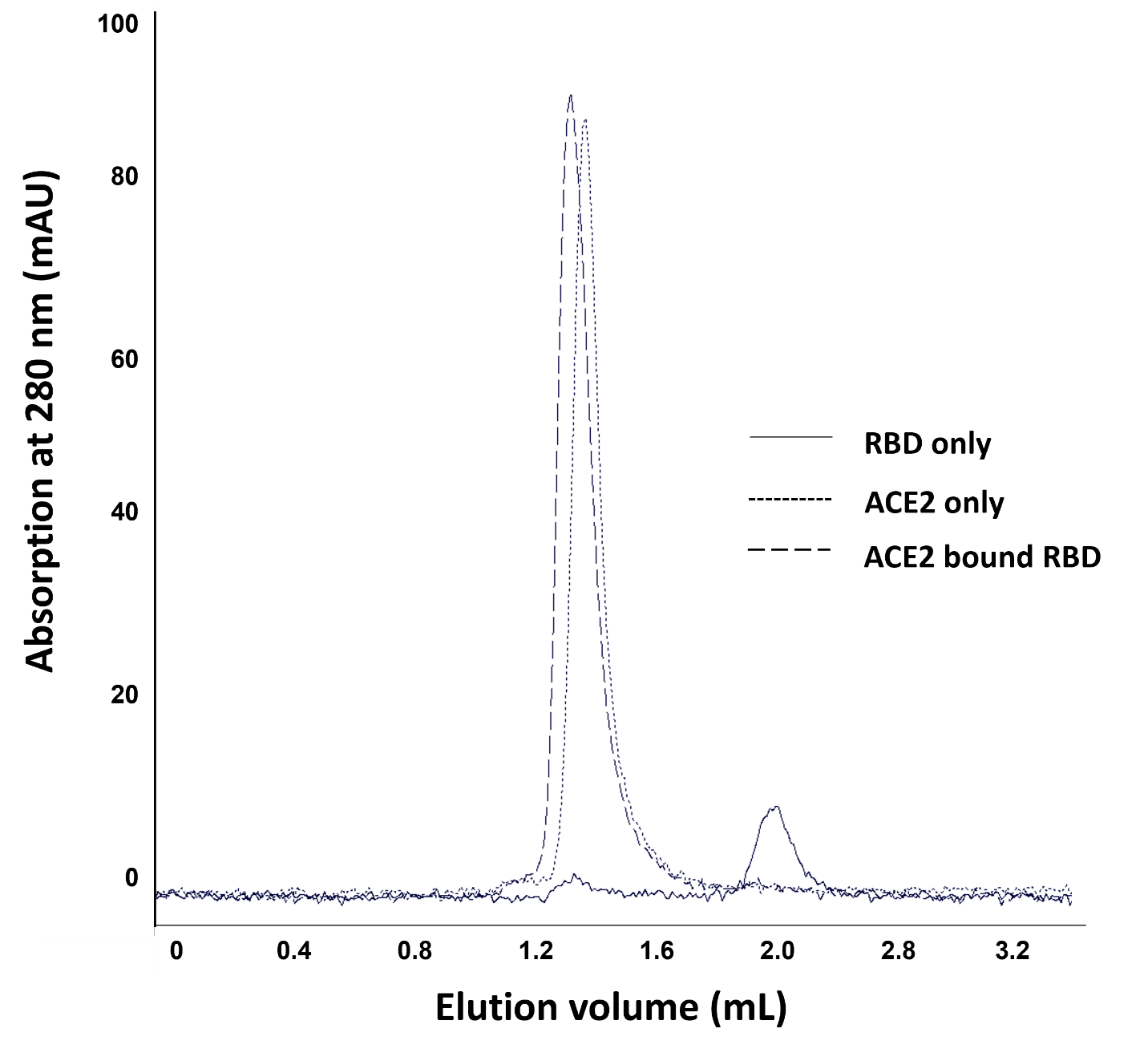

**Figure S3. Expression and purification of biotinylated SARS-CoV-2 RBD BA.4/5 variant protein.** A) Amino acid sequence of the BA.4/5 RBD protein. The signal peptide, Avi-tag, TEV protease site, and the hexa histidine-tag are underlined. B) Streptavidin gel shift assay was done in 12% SDS-PAGE gel and 981% of protein was biotinylated based on densitometry (ImageJ) analysis. Lane 1, BA.4/5 RBD only (2.5 µM). Lane 2, BA.4/5 RBD (2.5 µM) with Streptavidin (8.3 µM). Lane 3, Streptavidin only (8.3 µM). C) BA.4/5 RBD was purified by size exclusion chromatography (SEC) using a Superdex 75 16/600 column (Cytiva). Fractions B4 – C5 of the main peak (identified by black arrow) were collected. D) The purity of BA.4/5 RBD protein was determined by 12% SDS-PAGE gel after FPLC chromatography. E) The ACE2 binding of BA.4/5 RBD was evaluated by adding molar excess of ACE2 to RBD and analyzing the mixture by SEC using Superdex 200 Increase 5/150 column (Cytiva). By comparing the peak areas of RBD (~2.1 mL elution volume), we found that 94% of BA.4/5 RBD was bound to ACE2.

MGWSCIILFLVATATGVHSRVQPTESIVRFPNITNLCPFGEVFNATRFASVYAWNRKRISNCVADYSVLYNSASFSTFKCYGVSPTKLNDLCFTNVYADSFVIRGNEVSQIAPGQTGNIADYNYKLPDDFTGCVIAWNSNKLDSKVGGNYNYRYRLFRKSNLKPFERDISTEIYQAGNKPCNGVAGVNCYFPLQSYGFRPTYGVGHQPYRVVVLSFELLHAPATVCGPKKSTNLVKNKSVNFGGSGLNDIFEAQKIEWHEGGSENLYFQGGSGGGGSGGGGSSQSTIEEQAKTFLDKFNHEAEDLFYQSSLASWNYNTNITEENVQNMNNAGDKWSAFLKEQSTLAQMYPLQEIQNLTVKLQLQALQQNGSSVLSEDKSKRLNHHHHHH

B.

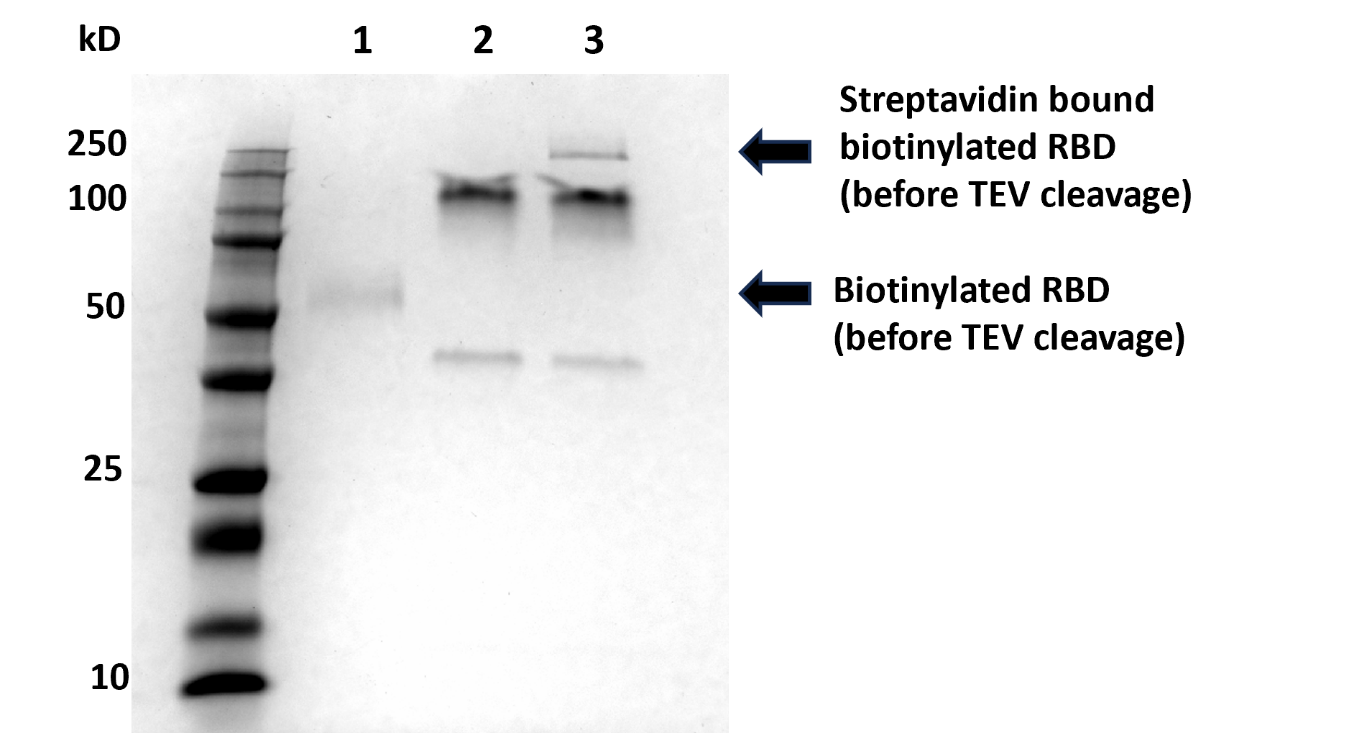

C.

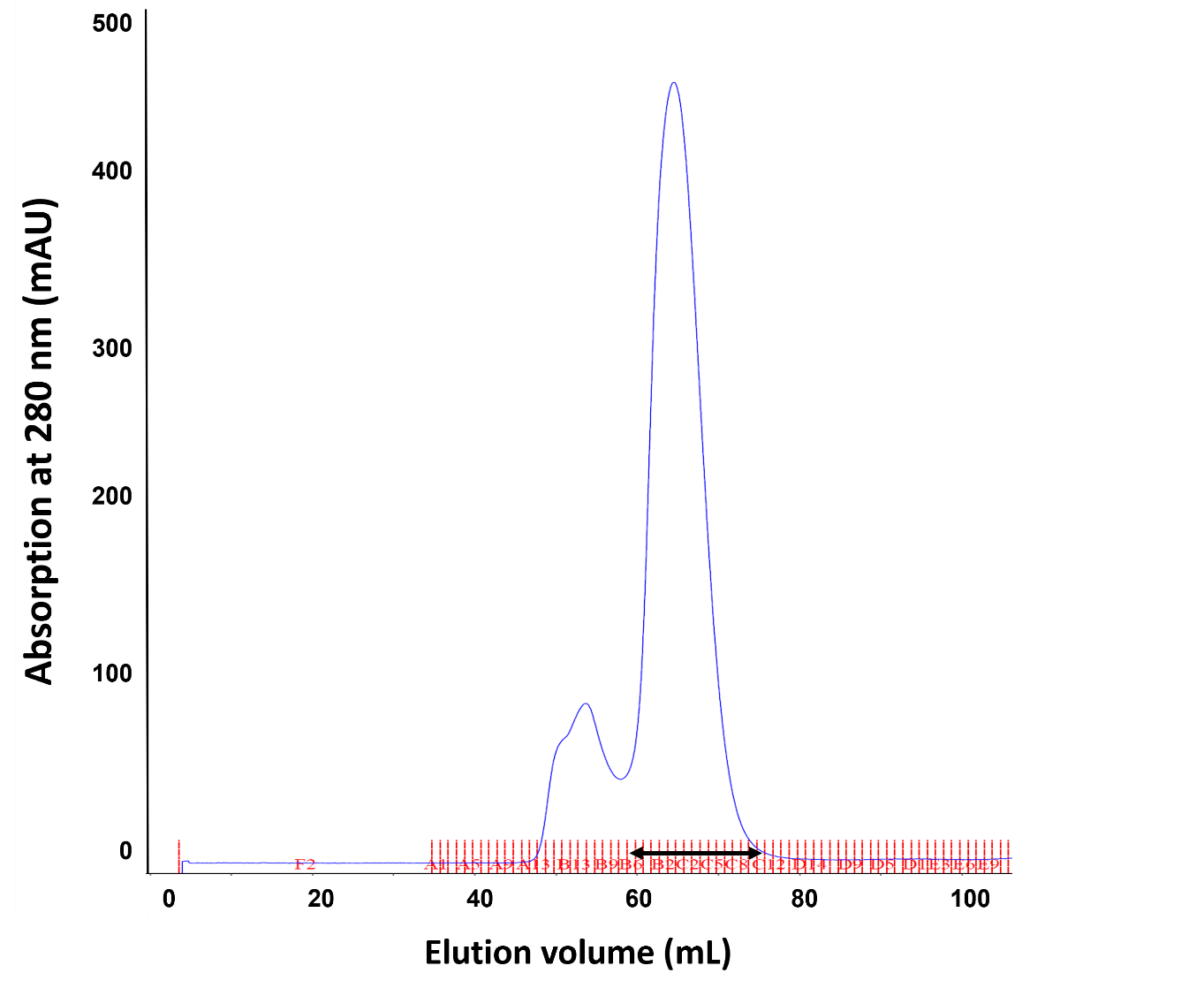

D.

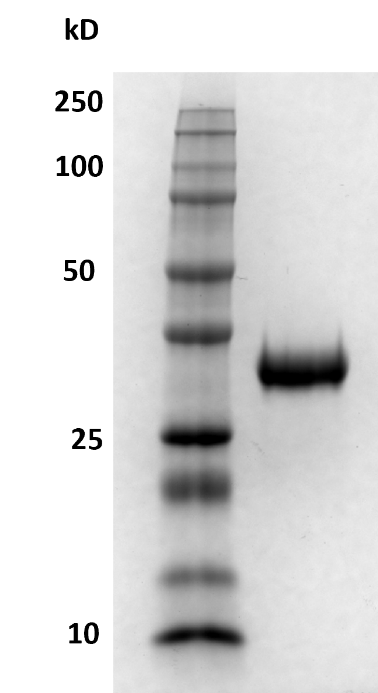

E.

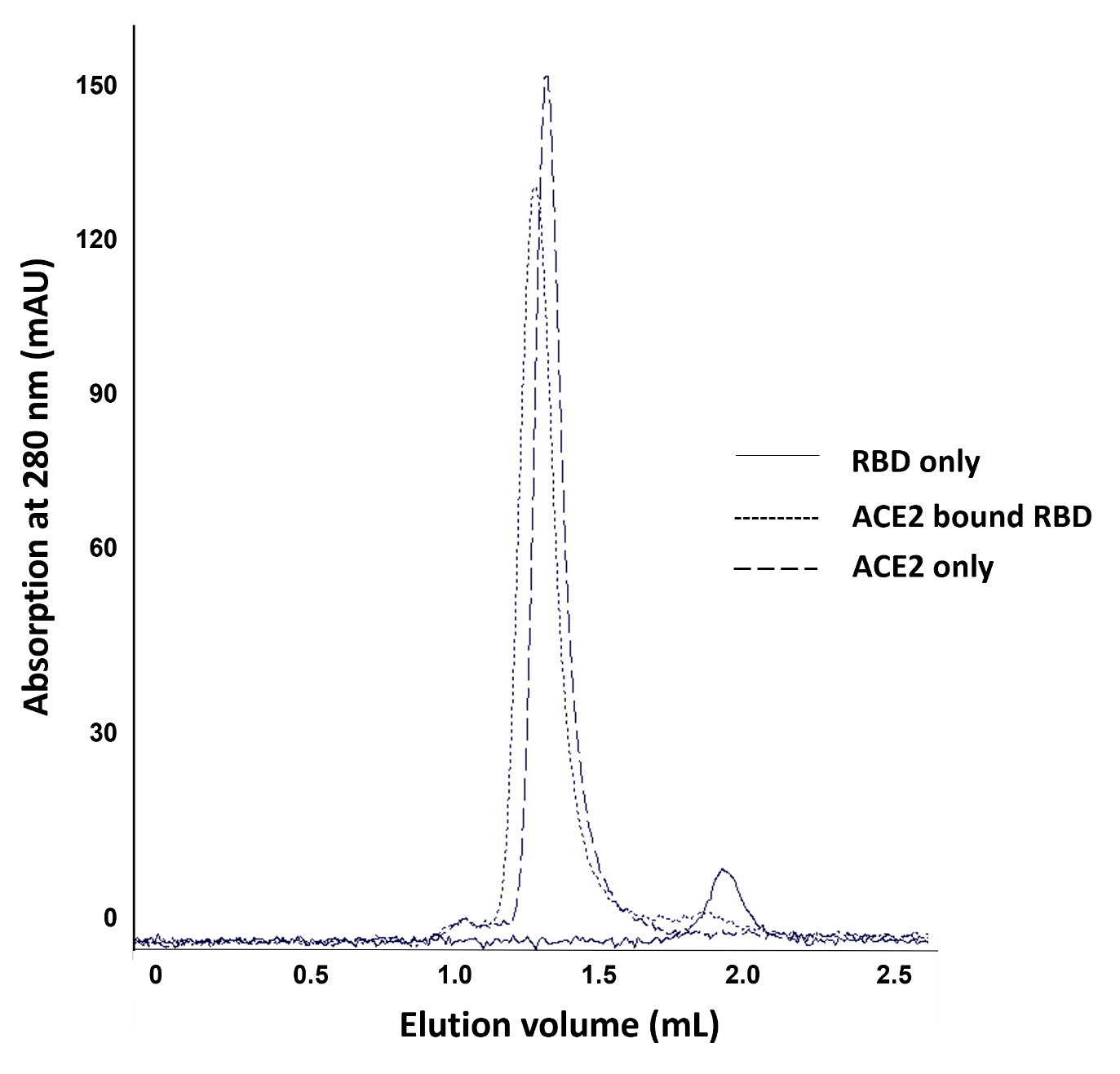

**Table S1. Results of interference screening in Q-NAb IgG Kit (Ancestral and BA.4/5).**

| **Category** | **Interferent** | **Concentration Tested** | **Ratio to control** | |
| --- | --- | --- | --- | --- |
|  |  |  | **Ancestral** | **BA.4/5** |
| Endogenous | Hemoglobin | 500 mg/dL | 103% | 113% |
|  | Bilirubin, conjugated | 40 mg/dL | 99% | 105% |
|  | Bilirubin, unconjugated | 40 mg/dL | 102% | 103% |
|  | Human Serum Albumin | 6 g/dL | 98% | 101% |
|  | Human IgA | 1.6 g/dL | 98% | 107% |
|  | Triglycerides | 1.6 g/dL | 94% | 109% |
|  | Biotin | 1200 ng/mL | 101% | 106% |
| Analgesic/  Fever reducer | Acetaminophen | 1324 uM | 110% | 109% |
|  | Ibuprofen | 2425 uM | 100% | 110% |
|  | Aspirin | 3.62 mM | 107% | 105% |
